# Contrastive Multi-modal Training with Electrocardiography and Natural Language Echocardiography Reports for Zero-shot Prediction of Structural Heart Disease

**DOI:** 10.1101/2025.09.16.25335870

**Authors:** Wai-Chak Wong, Che Liu, Pierre Elias, John Weston Hughes, Chun-Yu Leung, Xiao-Yan Qian, Hang-Long Li, Yuk-Ming Lau, Chao-Fan Tao, Ali Choo, Chi-Hang Yung, Chi-Hong Fong, Wai-Kwok Choi, Chak-Kong Cheng, Lok-Lam Cheng, Lik-Man Lau, Roshan Relwani, Jing Qin, Lequan Yu, Hin-Wai Lui, Ho-On Alston Conrad Chiu, Hung-Fat Tse, Chung-Wah Siu, Rossella Arcucci, Joshua Wing-Kei Ho, Chun-Ka Wong

## Abstract

**Background:** Machine learning models for predicting structural heart disease (SHD) from electrocardiography (ECG) traditionally required structured echocardiographic data. The potential of echocardiography (ECHO) natural language reports remains underused. We describe MERL-ECHO, a multimodal model using contrastive language-image pre-training (CLIP) that aligns ECG with ECHO natural language reports for zero-shot SHD prediction.

**Methods:** We conducted a multi-center retrospective study using paired ECG and ECHO natural language reports from Queen Mary Hospital and Tung Wah Hospital in Hong Kong. MERL-ECHO was trained on 45,016 pairs ECG-ECHO pairs. Performance was evaluated on an internal test set covering 10 SHDs and on an external test set of 5,442 ECGs with ECHO-derived labels for 6 SHDs from Columbia University Irving Medical Center, USA.

**Results:** The cohort included 8,192 patients (mean age 73.7±16.5 years; 55.3% male). In the internal test set, MERL-ECHO achieved an average AUROC of 0.69, with strongest performance for left ventricular dilation (0.78), right ventricular systolic dysfunction (0.71), and tricuspid regurgitation (0.71). In the external test set, the average AUROC was 0.72, with highest performance for left ventricular systolic dysfunction (0.76) and aortic stenosis (0.76). Pre-training improved AUROC by up to 5%, performance scaled with larger datasets, and ResNet18 outperformed ViT-Tiny as ECG encoder by 7%. Saliency analysis revealed interpretable ECG features, including unexpected P-wave changes in aortic stenosis, suggesting novel disease markers.

**Conclusions:** MERL-ECHO leverages ECHO natural language reports for multimodal training with ECG. This CLIP-based model enables accurate zero-shot prediction of SHDs and highlights interpretable ECG features with potential clinical relevance.

## INTRODUCTION

The global burden of heart failure due to valvular heart disease and impaired cardiac function has steadily increased over the past decades ^1–3^. Early detection of structural heart disease (SHD) is critical for enabling timely initiation of pharmacological and interventional therapies ^4–7^. Recent advances have demonstrated that deep neural networks (DNNs) applied to 12-lead electrocardiograms (ECGs) can accurately predict valvular heart disease ^8–10^.

The predictive accuracy of DNN models typically improves with larger training datasets ^11^. However, most prior models have relied on supervised learning with convolutional neural networks (CNNs) or recurrent neural networks (RNNs), which require structured datasets with ground-truth SHD severity labels. In clinical practice, echocardiography (ECHO) data may be captured as natural language free-text reports, leaving a vast amount of valuable information underutilized for ECG classifier training.

In the field of image classification, contrastive language-image pre-training (CLIP) has emerged as a groundbreaking self-supervised learning paradigm. This method enables training of image classifiers from paired images and natural language text without the need for manual labeling^12^. By leveraging large and diverse datasets, CLIP substantially expands the scope of model development. Building on this concept, we recently introduced Multimodal ECG Representation Learning (MERL), a model that applies the CLIP architecture for self-supervised training of ECG classifiers using paired ECG signals and natural language ECG reports ^13^. We hypothesize that MERL can be extended to incorporate paired ECG signals and natural language ECHO reports for the prediction of SHD.

In this multi-center study, we present MERL-ECHO, a CLIP-based model trained using contrastive multimodal learning on paired ECG signals and natural language ECHO reports. We demonstrate that MERL-ECHO enables zero-shot prediction of SHD across internal and external test sets. Furthermore, we systematically evaluate different architectural designs and report comparative results to guide future research in multimodal ECG-based SHD prediction.

## METHODS

### Study Design

This was a multi-center retrospective study involving patients from Queen Mary Hospital and Tung Wah Hospital, Hong Kong. This study conformed to the Declaration of Helsinki. Follows CONSORT-AI (Consolidated Standards of Reporting Trials-Artificial Intelligence) checklist for reporting ^14^. Consent from individual patients was waived as only anonymized data from the hospital registries was involved. Ethical approval was granted by HKU/HA HKW Institutional Review Board (HKU/HA HKW IRB; UW 25-042).

### Data Source

Adult patients aged 18 years or above having undergone 12-lead ECGs and echocardiograms at Queen Mary Hospital and Tung Wah Hospital, Hong Kong, from January 2008 to May 2025 were identified. ECGs were grayscale for the period 2008-2023 and coloured for the period 2023-2025. Natural language ECHO text reports for 2008-2025 were acquired. ECGs and natural language ECHO text reports were paired up. Only data pairs with the time difference between the ECG acquisition date and the ECHO text report date less than or equal to three years were included in the final study cohort. Each paired data was randomly assigned to either the training, validation, or test set (Figure 1).

**Figure 1.**
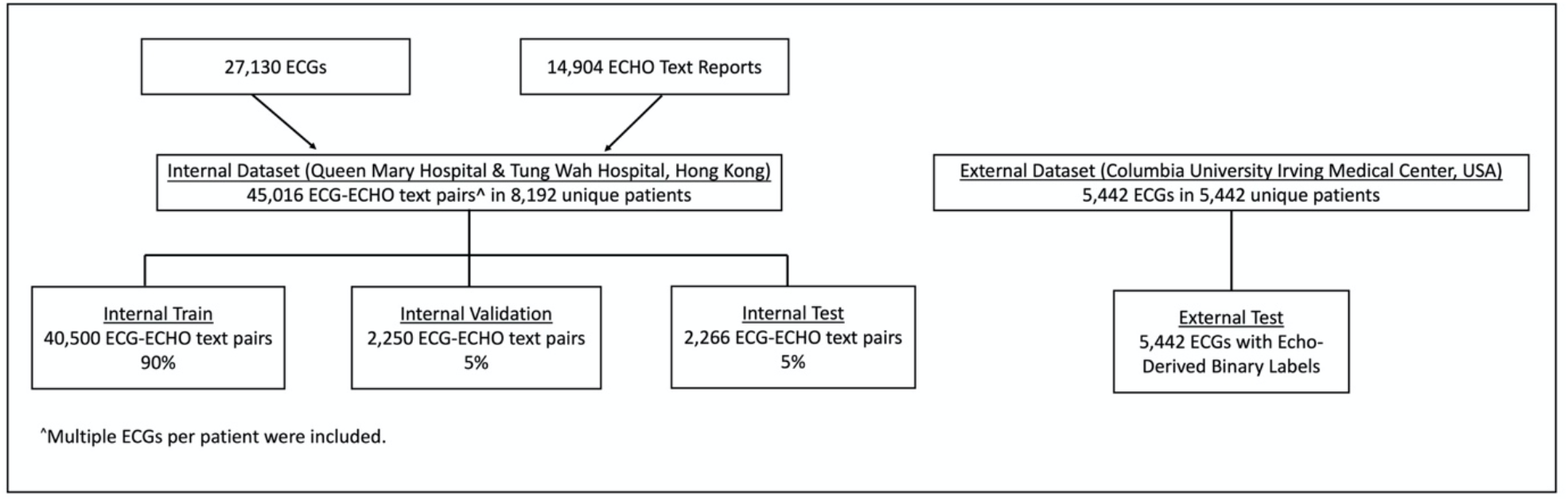
Patient Flow.

For the external test data set at Columbia University Irving Medical Center, all data were collected from adult patients aged 18 or above who had a digitally stored 12-lead ECG and a transthoracic echocardiogram within a 1-year interval between 2008 and 2022 (Figure 1). ECG waveform data were extracted from the GE MUSE ECG management system at a sampling frequency of 250 Hz across all 12 leads. Echocardiographic data were extracted from the Syngo Dynamics (Siemens) and Xcelera (Philips) systems. Each ECG was accompanied by the echo-derived binary labels, which were derived from structured echocardiogram reports and binarized using clinically relevant thresholds to indicate at least moderate disease severity.

### ECG pre-processing

Our training data were stored as images rather than raw digital signals. We utilized a simplified version of our previously developed DigitHeart pipeline to extract voltage-time series data for downstream use ^15^. First, a Yolo v11 object detection model was used to locate and label each of the 12 leads in ECGs ^16^ (Supplemental Figure S1), 99.5% of ECGs were successfully detected and cropped to the 12-lead region; the remaining ECGs (0.479%) were manually annotated to identify the 12 leads. Second, binary thresholding using OpenCV v4.10.0 with a cut-off value of 80 was used to grossly remove grid lines, followed by using connected components size filtering to further remove residual smaller grid lines ^17^. Third, two-dimensional binary images were converted into one-dimensional digital signals. The conversion algorithm raster-scanned the binary image matrix along its horizontal axis, which serves as the temporal domain, while the vertical axis represents the uncalibrated amplitude. For each pixel column (a discrete time step), the vertical centroid of the white pixels (representing the ECG waveform) was computed by averaging their y-coordinates. This centroid value represented a single sample point in the raw time series. If no white pixels were found in a column, the vertical position was interpolated by carrying forward the value from the preceding time step to maintain signal continuity. To reduce noise introduced during the image-to-signal conversion, the extracted signal was smoothed using a convolution-based moving average filter with a fixed window size of 10 applied. Finally, the extracted ECG waveform data in the internal dataset and ECG waveform data in the external dataset were up-sampled to a frequency of 500 Hz and stored in .mat format. Upsampling the digital signal to 500 Hz ensures that the fine-tuned data inputs are consistent with the pre-training ECG data during the pre-training stage of MERL-ECHO, thereby minimizing potential distribution shift arising from disparate sampling rates. To meet the model’s input requirement of 10-second ECG recordings sampled at 500 Hz, the up-sampled ECG waveforms were repeated as needed to achieve the required duration.

### ECHO natural language text report pre-processing

First, we used the Qwen3-4B model, a general-purpose large language model, to analyze the natural language text of each ECHO report ^18^. Our objective was to exclude ECG–ECHO text pairs whose reports documented any cardiac valve interventions listed in Supplemental Table S1; only pairs linked to reports without these interventions were included. Second, the cardiac abbreviation terms were expanded to the full terms, such as “AS” to “aortic stenosis” and “MR” to “mitral regurgitation”. Details are provided in the Supplemental Table S2. Empty or ECHO reports with less than 4 words were excluded. This text pre-processing preserves the ECHO text in a form that is as close as possible to its original state. We will also explore whether imposing a more structured representation of ECHO text yields different results; the structured pre-processing approach will be discussed in a subsequent section.

### Diagnosis annotation

To validate the performance of our MERL-ECHO model, we developed a web application for annotating a subset of our internal data sets to serve as ground truth (Supplemental Figure S2). A committee consisting of 3 cardiologists and trained personnel was responsible for annotating 10 types of labels for each ECHO report, including aortic, mitral, and tricuspid stenosis and regurgitation, left and right ventricular dilation and systolic impairment, and left ventricular hypertrophy based on the same thresholding criteria. The first 500 ECHO reports were manually annotated. Thereafter, the remaining 4,016 ECHO reports were annotated using a semi-automated, human-in-the-loop workflow. In this pipeline, locally deployed large language model Qwen3-4B was used to generate initial predictions for each report (average F1 score 0.77; Supplemental Figure S3; Supplemental Table S3) ^18^. Subsequently, all reports were reviewed and corrected by human annotators to ensure the final accuracy and fidelity of all labels. This hybrid approach significantly accelerated the data curation process while maintaining expert-level quality.

### CLIP model development and training

Our methodology is predicated on a multimodal learning framework designed to train a zero-shot diagnosis predictor, MERL-ECHO, to predict SHDs from ECGs by integrating ECG signals with their corresponding natural language ECHO text reports. This approach, inspired by the principles of CLIP and adapted for the medical signal-text domain ^12^, leverages the rich semantic information contained within clinical narratives to inform and structure the learned ECG representations. We previously described the use of CLIP architecture to train a zero-shot ECG diagnosis predictor (MERL) by using ECG signal and ECG text report pairs ^13^. In this study, we want to detect SHDs from ECGs by training a model (MERL-ECHO) using paired ECGs and natural language ECHO text reports.

MERL-ECHO is a dual-encoder architecture, featuring two distinct encoders for ECG signals and ECHO text reports, which are used to process the corresponding modalities, respectively. During the training process, all parameters in the ECG encoder will be updated, while only those in the last 6 layers of the text encoder will be updated. In our whole dataset 𝒳, we represent each ECG-ECHO text pair as (e_i_, r_i_), where as e_i_ ∈ ε denotes the raw ECG signal and r_i_ ∈ ℛ denotes the associated natural language ECHO text report, respectively, with *i* = 1, 2, 3, …, *N*. Two contrastive learning strategies are utilized and described as follows.

The first and most important is the Cross-Modal Alignment (CMA). It applied a contrastive learning approach on ECG-ECHO text pairs (Figure 2A) and enables MERL-ECHO to align the feature representation of an ECG signal with the feature representation of its corresponding natural language ECHO text report. ECG encoder with 1D-ResNet18 ^19^ or ViT-Tiny ^20^ as the backbone embeds the ECG signal, and Med-CPT ^21^ as the text encoder embeds the natural language ECHO text report. Each ECG-ECHO text pair (e_i_, r_i_) will be embedded into the shared, high-dimensional latent embedding space, denoted as (*z*_*e,i*_, *z*_*r,i*_), by the corresponding encoders. Subsequently, the ECG embedding and ECHO-text embedding will be mapped into the same dimensionality *d* by two different non-linear projectors (*P*_*e*_ and *P*_*r*_), they are represented as 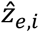 and 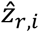, respectively. Third, cosine similarities for ECG-ECHO text and ECHO Text-ECG will be computed as 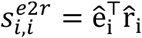 and 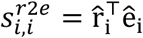, respectively. 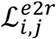 and 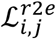 represent ECG-ECHO text and ECHO text-ECG cross-modal contrastive losses, the final loss function (ℒ_*CMA*_) in cross-modal alignment is the average of these two losses (Formula 1), allowing MERL-ECHO to be trained to maximize the cosine similarity of embeddings from true pairs while minimizing the cosine similarity of non-paired embeddings, and making the alignment learning robust and bidirectional:

**Figure 2.**
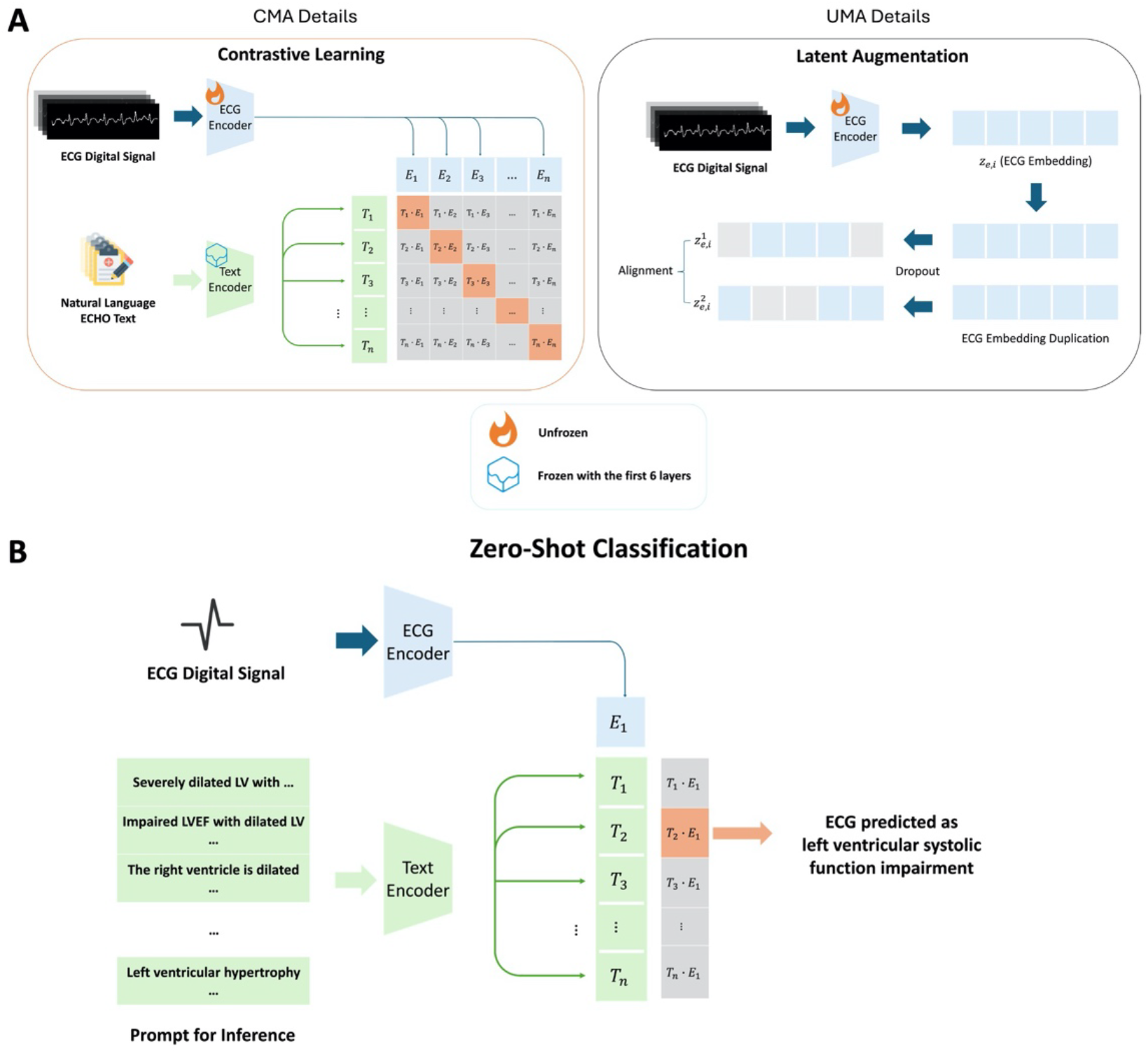
Model Architecture and Zero-shot Capability.

**Formula 1:**

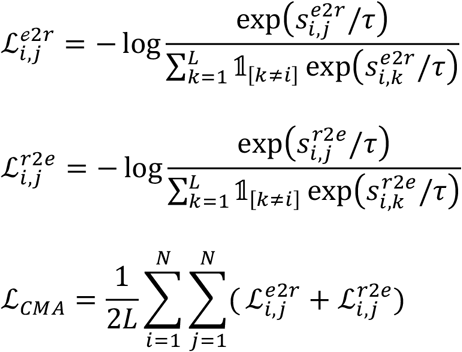

Cross-Modal Alignment strategy not only aligns ECG signal features and semantic features from natural language ECHO text report, but it also enables MERL-ECHO the capability of zero-shot classification (Figure 2B). As ECG and ECHO-text features are aligned in the same high-dimensional latent embedding space during the training process, ECG signal features are injected with high-level, human-understandable semantic information from ECHO-text. We can test the model by simply providing a text prompt and an ECG signal. The model can then find matching ECGs by calculating feature similarity, without requiring any labelled data or fine-tuning for the downstream task.

In addition to the alignment between the features of ECG and ECHO-text, we further employ Uni-Modal Alignment (UMA). It is also a contrastive learning method, but it operates within a single modality, specifically the ECG domain (Figure 2A). Its purpose is to learn higher-quality, more discriminative feature representations for ECGs. There are three steps in this workflow: First, the ECG encoder embeds the ECG signal to obtain *z*_*e,i*_. Then, embedding *z*_*e,i*_ is duplicated. Independent and random dropout with a probability (*p*) 0.1 is applied to these two identical embeddings for generating the positive pair 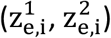 ^22^ (Supplemental Figure S4 for notation details). Third, we use standard contrastive loss on the positive pair and treat other unpaired combinations as negative pairs (Formula 2). This latent augmentation technique, by using two independent dropout operations to construct positive pairs, allows MERL-ECHO to learn ECG signal features that are more powerfully discriminative. It works in tandem with CMA (Cross-Modal Alignment) to form the training core of the MERL-ECHO framework.

**Formula 2:**

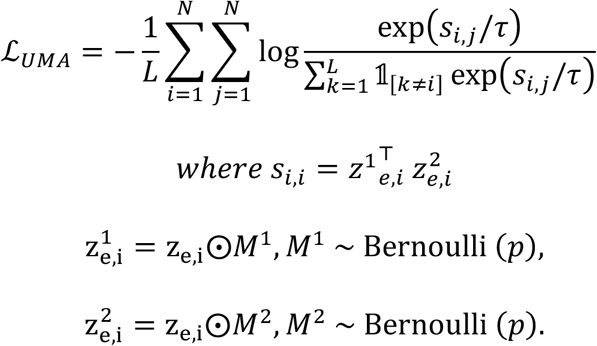

To further enhance performance, we also included a pre-training phase based on the previous work from our group ^13^, where a CLIP-based model was trained on 771,693 ECG digital signals and ECG text report pairs initially on the publicly accessible dataset, MIMIC-ECG ^23^, and it enabled zero-shot classification for various electrocardiographic abnormalities, we leverage pre-trained model weights to facilitate knowledge transfer (Supplemental Figure S5) from ECG-based representations to echocardiographic insights.

We trained the model for 15 epochs on the training set with the AdamW optimizer, setting a learning rate of 1 × 10^−5^ and a weight decay of 1 × 10^−8^. We unfroze the last six layers of the text encoder. We applied a cosine annealing scheduler for learning rate adjustments and maintained a batch size of 256 per GPU (two GeForce RTX 4090D). We set up early stopping criteria if the evaluation loss did not improve for three consecutive epochs.

### Prompts for zero-shot classification during inference

During inference, a key feature of our CLIP-based model is the capability to perform zero-shot classification. Our model can predict any arbitrary diagnosis from a new ECG by inputting text prompts, without the need for retraining (Figure 2B). By directly leveraging ECG waveform signals along with carefully engineered text prompts for 10 distinct echocardiographic prediction classes, the model can predict the binary classification for each of the echocardiographic labels for the ECG by calculating the cosine similarity between the ECG embedding and each text prompt embedding. The ECG-ECHO text pair with the highest similarity will be the output.

To prevent model overfitting to the text prompt during the training stage, we utilized the diagnostic category names as the text prompts. Prompts for zero-shot inference were explicitly designed to enhance model prediction performance.

A prompt is a natural language text describing the semantic context of an image or a task. By designing appropriate prompt engineering, the CLIP-based model can better align the echocardiographic textual description, the prompt, with the ECG digital signal. To optimize prompt design, we introduced enhanced cardiology domain prompt engineering to allow prompts with variant phrases and associated features. The complete prompt list is in Supplemental Table S4. We first queried a large language model, GPT-4, which has been shown to achieve high accuracy in generating cardiologist-level ECHO reports ^24–27^, to generate prompts for 10 echocardiographic labels. Query details are in Supplemental Table S5. Following prompt generation, experienced cardiologists reviewed and refined the prompts based on accuracy, clinical relevance, and clarity criteria. This multi-step validation ensures that the generated prompts accurately reflect the underlying cardiac pathophysiology and are contextually appropriate for domain-specific zero-shot classification tasks.

### Saliency Mapping

To better understand the ECG representations that are most important to model prediction, we used the Gradient-weighted Class Activation Mapping (Grad-CAM) technique to visualize ECGs with positive classes and the highest confidence model predictions ^28^. To do this, our model first receives both ECG signals and text prompts simultaneously, extracting features through the ECG encoder and text encoder, respectively, and calculating similarity scores via dot products, which serves as the target output of Grad-CAM analysis. To generate the saliency map, we targeted the final convolutional layer within the last residual block of our ECG encoder. During the backward propagation, the gradients of the similarity score with respect to the feature maps of this target layer were computed. These gradients were then global-average-pooled to obtain a set of weights, representing the importance of each feature map. A weighted linear combination of the feature maps was computed using these weights, followed by a rectified linear unit (ReLU) activation to isolate features with a positive influence on the prediction. The resulting saliency map is then up-sampled to the original length of 5000 time steps using linear interpolation. This global saliency map is further overlaid onto each of the 12 leads of the original ECG waveform. It provides a comprehensive view, where the heat intensity (color-coded in red) indicates the temporal importance, as determined by the model, allowing for a qualitative assessment of which segments of the cardiac cycle contribute most significantly to the final similarity score.

### Statistical Analysis

Descriptive statistics using counts, percentages, and medians were provided as appropriate. Discrete variables were presented as mean ± standard deviation. F1 score, area under the receiver operating curves (AUROC), and area under the precision-recall curves (AUPRC) were calculated for the model prediction on all classes in both the internal and external test sets to assess their performance. The F1 score was calculated as the harmonic mean of the precision and recall using a method that achieved the best balance between precision and recall; a series of thresholds were calculated to get different values of precision and recall, and the F1 score was chosen when the calculated precision and recall resulting the maximum F1 score based on the specific threshold value that was also the cut-off value used to distinguish between positive and negative class. It helps the model to find an optimal classification threshold that achieved the best predictive performance (measured by the F1 score). The range of F1 score values is from 0 to 1, where 1 indicates perfect classification performance, and 0 indicates the worst possible performance. The AUROC was used to describe model accuracy, plotting sensitivity (true positive rate) against 1-specificity (false positive rate) with values ranging from 0.5 to 1, where 1 indicates perfect classification, and 0.5 indicates random guessing. The AUPRC was also used to describe model accuracy, plotting precision (positive predictive value) against recall (sensitivity). All statistical analyses were performed using Python 3.12.3.

## RESULTS

### Patient Characteristics

A total of 45,016 unique ECG signals and ECHO text pairs were identified in 8,192 unique patients from 2 public hospitals in Hong Kong, namely Queen Mary Hospital and Tung Wah Hospital, respectively, of whom 4530 (55.3 %) were male, and 3662 (44.7 %) were female. Their mean age was 73.7±16.5 years. Patient characteristics for the training, validation, test, and external test sets are described in **Table 1**.

**Table 1.** Patient Characteristics.

|  | Train<sup>#</sup> | Validation | Test | External test |
| --- | --- | --- | --- | --- |
| No. of ECG-ECHO Pairs | 40,500 | 2,250 | 2,266 | 5,442 |
| Patients, n | 7,099<sup>##</sup> | 1,693<sup>##</sup> | 1,638<sup>##</sup> | 5,442 |
| Age, mean±SD, y | 73.5±16.6 | 72.4±15.8 | 71.4±16.7 | 71.1±16.7<sup>*</sup> |
| Sex, % |  |  |  |  |
| Female | 3,206 (45.2%) | 764 (45.1%) | 731 (44.6%) | 2,731 (50.2%) |
| Male | 3,893 (54.8%) | 929 (54.9%) | 907 (55.4%) | 2,711 (49.8%) |
| Ventricular Size and Morphology, % |  |  |  |  |
| Left Ventricular Hypertrophy | 25.7 | 18.3 | 19.1 | - |
| Dilated Left Ventricle | 10.3 | 8.6 | 9.1 | - |
| Dilated Right Ventricle | 13.2 | 10.7 | 10.7 | - |
| Ventricular Function, % |  |  |  |  |
| Impaired Left Ventricular Systolic Function<br>(Moderate to Severe) | 46.6 | 43.5 | 44.3 | 17.7 |
| Impaired Right Ventricular Systolic Function<br>(Moderate to Severe) | 12.7 | 10.3 | 10.9 | 7.7 |
| Moderate to Severe Valvular Heart Diseases, % |  |  |  |  |
| Aortic Stenosis (AS) | 9.6 | 9.9 | 9.3 | 5.3 |
| Aortic Regurgitation (AR) | 20.8 | 10.2 | 9.7 | 1.2 |
| Mitral Stenosis (MS) | 12.1 | 11.6 | 12.3 | - |
| Mitral Regurgitation (MR) | 35.8 | 26.4 | 26.8 | 6.2 |
| Tricuspid Regurgitation (TR) | 30.0 | 28.4 | 27.8 | 6.5 |

#Multiple ECGs and ECHO text reports per patient were included to maximize model training.

### Model Performance

The model performance in AUROC and AUPRC of each echocardiographic finding label is shown in Figure 3 and Table 2. In the internal test set, the average AUROC is around 0.69. It varied slightly across categories and diseases (Figure 3A). For ventricular function, the model achieved the highest AUROC of 0.71 in right ventricular systolic function impairment (RV impaired). In the category of ventricular size, the highest AUROC was found for left ventricular dilation (LV dilated; 0.78). Among valvular heart diseases, moderate to severe tricuspid regurgitation had the highest AUROC of 0.71. And left ventricular hypertrophy had the lowest AUROC of 0.64. The AUROC values for moderate to severe aortic and mitral stenosis, as well as aortic and mitral regurgitation, were 0.67. For the area under the precision-recall curves and F1 score, MERL-ECHO achieved the highest AUPRC of 0.63 and the highest F1 score of 0.66 in left ventricular systolic function impairment (LV impaired) (Figure 3B).

**Table 2.** Diagnostic performance by AUROC and AUPRC using MERL-ECHO for each label.

| Echocardiographic<br>Finding Label** | Internal Test |  | External Test |  |
| --- | --- | --- | --- | --- |
|  | AUROC | AUPRC | AUROC | AUPRC |
| LV Dilated | 0.78 | 0.27 | - | - |
| LV Impaired | 0.70 | 0.63 | 0.76 | 0.48 |
| RV Dilated | 0.68 | 0.21 | - | - |
| RV Impaired | 0.71 | 0.20 | 0.71 | 0.17 |
| AS Mod Severe | 0.67 | 0.15 | 0.76 | 0.14 |
| AR Mod Severe | 0.67 | 0.15 | 0.69 | 0.02 |
| MS Mod Severe | 0.67 | 0.20 | - | - |
| MR Mod Severe | 0.67 | 0.42 | 0.69 | 0.13 |
| TR Mod Severe | 0.71 | 0.50 | 0.72 | 0.17 |
| LVH | 0.64 | 0.30 | - | - |

\*\* LV Dilated = Left Ventricular Dilation, LV Impaired = Left Ventricular Systolic Function Impairment, RV Dilated = Right Ventricular Dilation, RV Impaired = Right Ventricular Systolic Function Impairment, AS Mod Severe = Moderate to Severe Aortic Stenosis, AR Mod Severe = Moderate to Severe Aortic Regurgitation, MS Mod Severe = Moderate to Severe Mitral Stenosis, MR Mod Severe = Moderate to Severe Mitral Regurgitation, TR Mod Severe = Moderate to Severe Tricuspid Regurgitation, LVH = Left Ventricular Hypertrophy.

**Figure 3.**
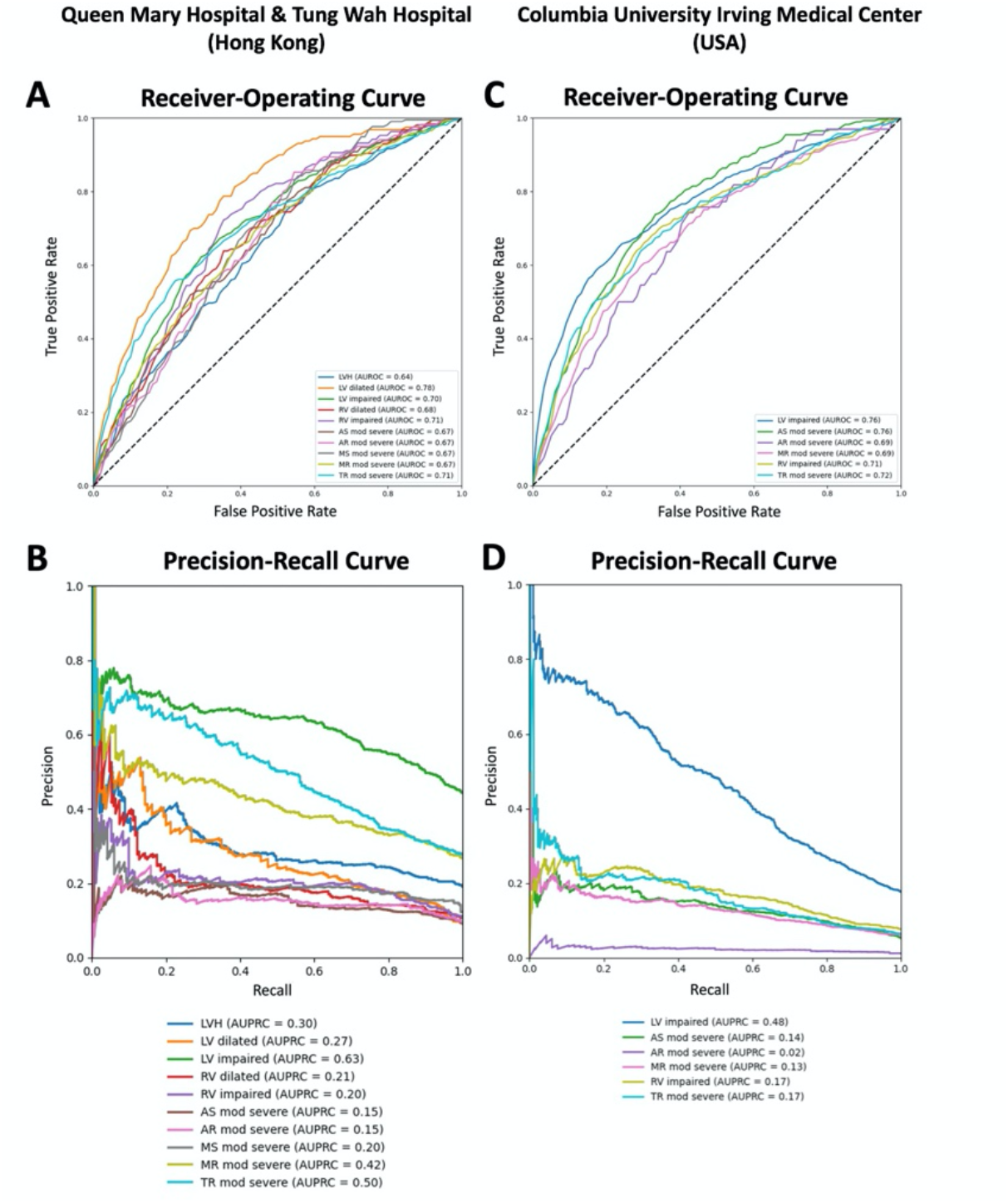
AUROC and AUPRC for the Internal and External Test Sets.

In the external test set (Columbia University Irving Medical Center), patients’ characteristics were similar to those in the internal data set in terms of sex and age (Table 1). Surprisingly, MERL-ECHO even performs better in the external test set than in the internal test. The average AUROC for 6 echocardiographic classification labels is 0.72 (Figure 3C). The AUROC for left ventricular systolic function impairment and moderate to severe aortic stenosis was the highest (AUROC: 0.76), and the lowest AUROC was found for moderate to severe aortic regurgitation (AUROC: 0.68). For the area under the precision-recall curves and F1 score, MERL-ECHO achieved the highest AUPRC of 0.48 and the highest F1 score of 0.49 in left ventricular systolic function impairment (LV impaired) (Figure 3D). Although there were variations in the AUROC for some echocardiographic labels, the model showed its ability to be robust and generalizable to the external data.

### ECG Encoder

For these two ECG feature extractor networks, the CNN-based ResNet18 and transformer-based ViT-Tiny, the zero-shot performance using different data proportions and pre-training on ECG-ECG text data shows that our CLIP-based model with CNN-based ResNet18 generally surpasses ViT-Tiny’s (Figure 4A). The ECG encoder using ResNet18 as the backbone surpasses the ViT-Tiny one by almost 7% in AUROC when using the whole training dataset (Table 3).

**Table 3.** AUROC comparison for two ECG encoders under different training data sizes.

| Metrics | Pretraining<sup>###</sup> | ECG Encoder | Training Set Size |  |  |  |  |
| --- | --- | --- | --- | --- | --- | --- | --- |
|  |  |  | 8,100 (20%) | 16,200 (40%) | 24,300 (60%) | 32,400 (80%) | 40,500 (100%) |
| AUROC | ✓ | ResNet | 59.9 | 63.1 | 64.3 | 66.9 | 68.9 |
|  |  | ViT | 51.7 | 56.5 | 58.2 | 60.2 | 62.7 |
|  | ✗ | ResNet | 49.4 | 56.0 | 57.4 | 59.7 | 63.6 |
|  |  | ViT | 52.8 | 51.8 | 53.1 | 52.2 | 52.2 |

<sup>###</sup>Pre-training with ECG-ECG text pair data.

**Figure 4.**
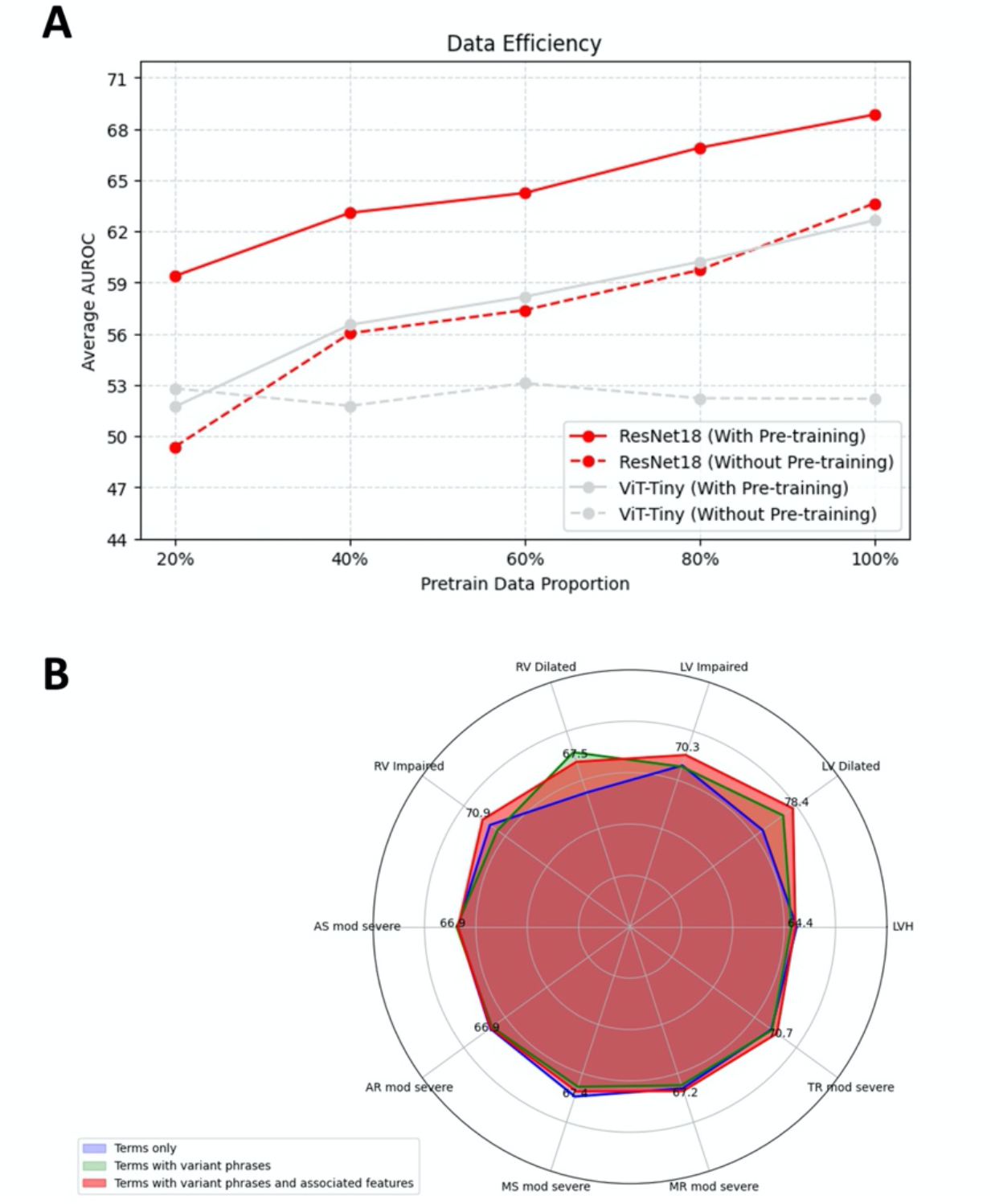
Data Efficiency Plot and Radar Plot for AUROC under Different Prompts.

### Effect of ECG-ECG Text Pre-training

Pre-training is the initial stage of the model training; it allows the model to learn general ECG features and patterns by initially training on a large amount of ECG-ECG text pair data without manually labelled ground-truth, and provides a foundation for subsequent fine-tuning. With pre-training on ECG-ECG text pair data, we observed that fine-tuning on the complete ECG-ECHO text training set led to improved model performance for ResNet18 as ECG encoders (Table 3), compared to models trained without pre-training. Specifically, for the CNN-based ResNet18 model, pre-training resulted in the highest AUROC of around 69% in the internal test set, representing a 5% improvement over the model without pre-training.

### Effect of Scaling Training Data

We also conducted comprehensive experiments to investigate the effect of scaling training data on AUROC. All the models with ResNet18 as the ECG encoder, with the pre-training and without the pre-training on ECG-ECG text data, improve AUROC steadily (Figure 4A). Crucially, the performance trend did not show signs of reaching a plateau, as AUROC continued to increase steadily up to the full dataset size. Specifically, with pre-training, the model’s performance with ResNet18 as ECG encoder rises by 9% in AUROC when using the whole training dataset compared to using 20% of data only (Table 3), and the ResNet-based encoder outperformed its transformer-based Vit-Tiny counterpart, achieving a 6% higher AUROC. This sustained upward trend suggests that there is still potential for increasing AUROC because of the continuously growing trend for AUROC.

### Ablation Study on Text Preprocessing

To evaluate the necessity of explicit feature engineering within the natural language clinical text, we conducted an experiment comparing two text-processing strategies. The first strategy involved training the model on natural language ECHO text reports, where only common abbreviations were expanded into their full terms. The second strategy involved changing the numerical values of the indicator into a specific diagnosis (Supplemental Table S6), making natural language in a more structured way. For instance, “LVEF = 45%” was transformed into “moderate left ventricular systolic function impaired”. Our results showed that providing explicit categorical labels for LVEF yielded no performance benefit. In fact, the model trained on unaltered, natural language text achieved a slightly superior AUROC. The model using natural language text reached an average AUROC of 0.72 in the external test set, marginally outperforming the model trained on categorized text, which scored an average AUROC of 0.71. Specifically, the model trained on natural language ECHO text achieved 0.76 AUROC in left ventricular systolic function impairment in the external test set, marginally outperforming the model trained on categorized text, which scored 0.75 AUROC in the same category. This finding suggests that the model is capable of learning the clinical significance of numerical LVEF values directly from the contextual information present in the raw text, rendering manual categorization unnecessary.

### Prompt Design Optimisation

In addition to the prompts generated by using our enhanced cardiology domain prompt engineering strategy, two more types of prompts, term-based prompts, and variant phrase prompts, were generated by GPT-4 to investigate the effects of prompts in zero-shot classification. Query details for the prompts generation are in Supplemental Table S5. Example prompts for left ventricular dilation and its variations are shown below:

1. **Terms only:** Consisting solely of the diagnostic category name.
  - Example: *“Dilated left ventricle*.*”*
2. **Terms with variant phrases:** Producing multiple phrasings or syntactic variants for each prediction class to capture linguistic diversity.
  - Example: *“Dilated left ventricle, LV dilation, LV enlargement (LVEDD > 58 mm in males or > 52 mm in females)*.*”*
3. **Terms with variant phrases and associated features:** Incorporating additional echocardiographic descriptive phrases that complement and do not contradict the definitions of the prediction classes.
  - Example: *“severely dilated LV with impaired LVEF at 30%, moderate mitral regurgitation (MR), and dilated left atrium (LA)*.*”*

Our prompt list is presented in Supplemental Table S4, with terms with variant phrases and associated features achieving the highest average AUROC of 0.69 among them (Figure 4B; Supplemental Table S5). The advantage of using prompts of terms with variant phrases and associated features is mainly demonstrated in the zero-shot inference of left ventricular dilation and systolic function impairment, and right ventricular systolic function impairment. We conclude that making the prompt more comprehensive and detailed may be helpful in improving accuracy.

### Saliency Mapping

To better understand which ECG features contributed most to model predictions, we applied Grad-CAM to visualize salient regions identified by the MERL-ECHO model. This approach helps address the interpretability challenges of black-box algorithms. Representative Grad-CAM images are shown in Figure 5. In the illustrative ECG predicted as high risk for left ventricular impairment in figure 5A, the model highlighted poor R-wave progression in the QRS complex and premature ventricular complexes as important features. These findings are consistent with established clinical knowledge ^29–31^.

**Figure 5.**
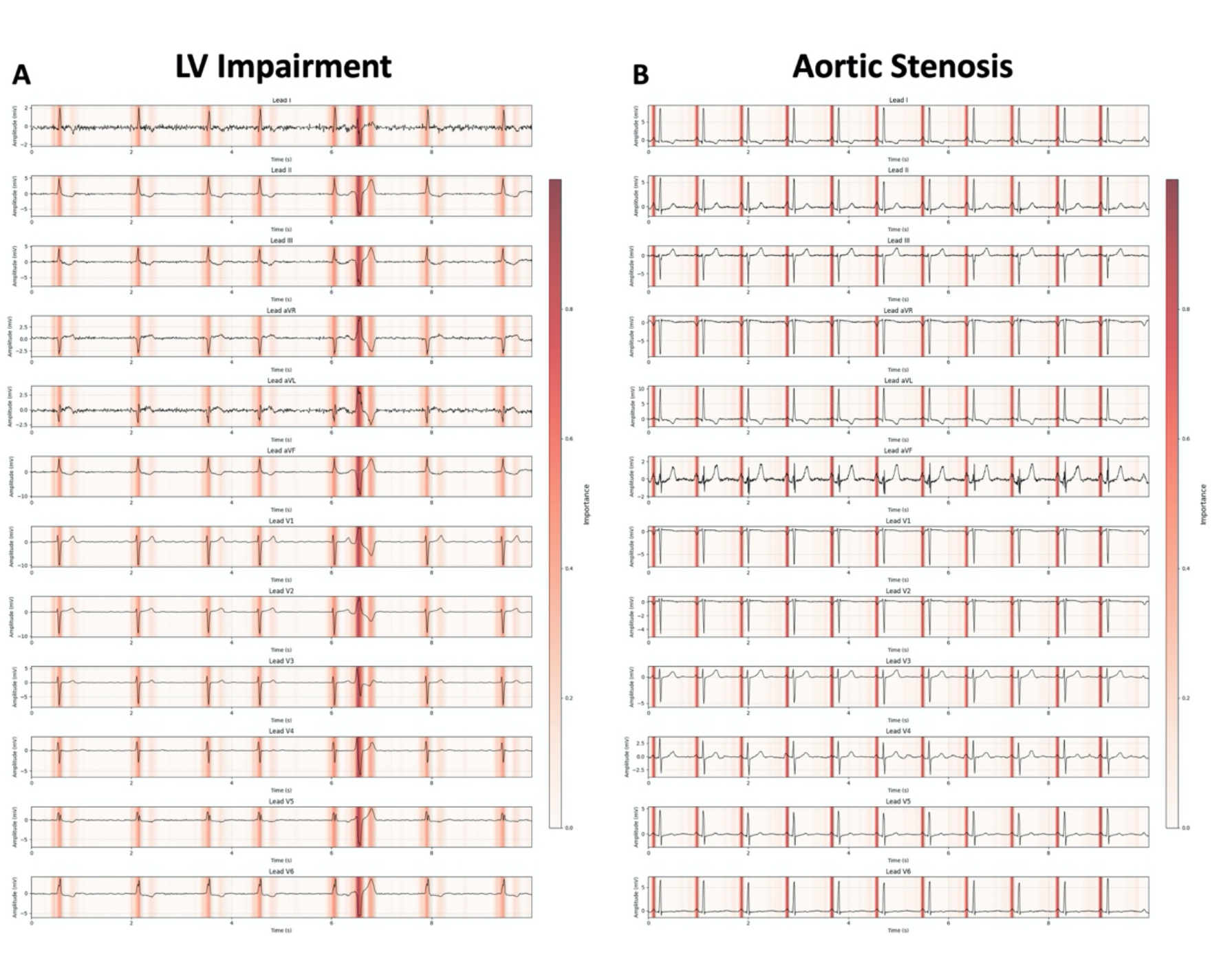
Saliency Map.

Moreover, MERL-ECHO consistently emphasized the P wave as the most important feature for predicting aortic stenosis (Figure 5B). This observation was unexpected and prompted further investigation. We therefore examined P-wave morphologies in patients with and without aortic stenosis (Supplemental Method S1). Quantitative analysis revealed that patients with aortic stenosis had a higher prevalence of flattened P waves in leads I, V5, and V6 (Figures 6A and 6B, and Supplemental Tables S7 and S8).

**Figure 6.**
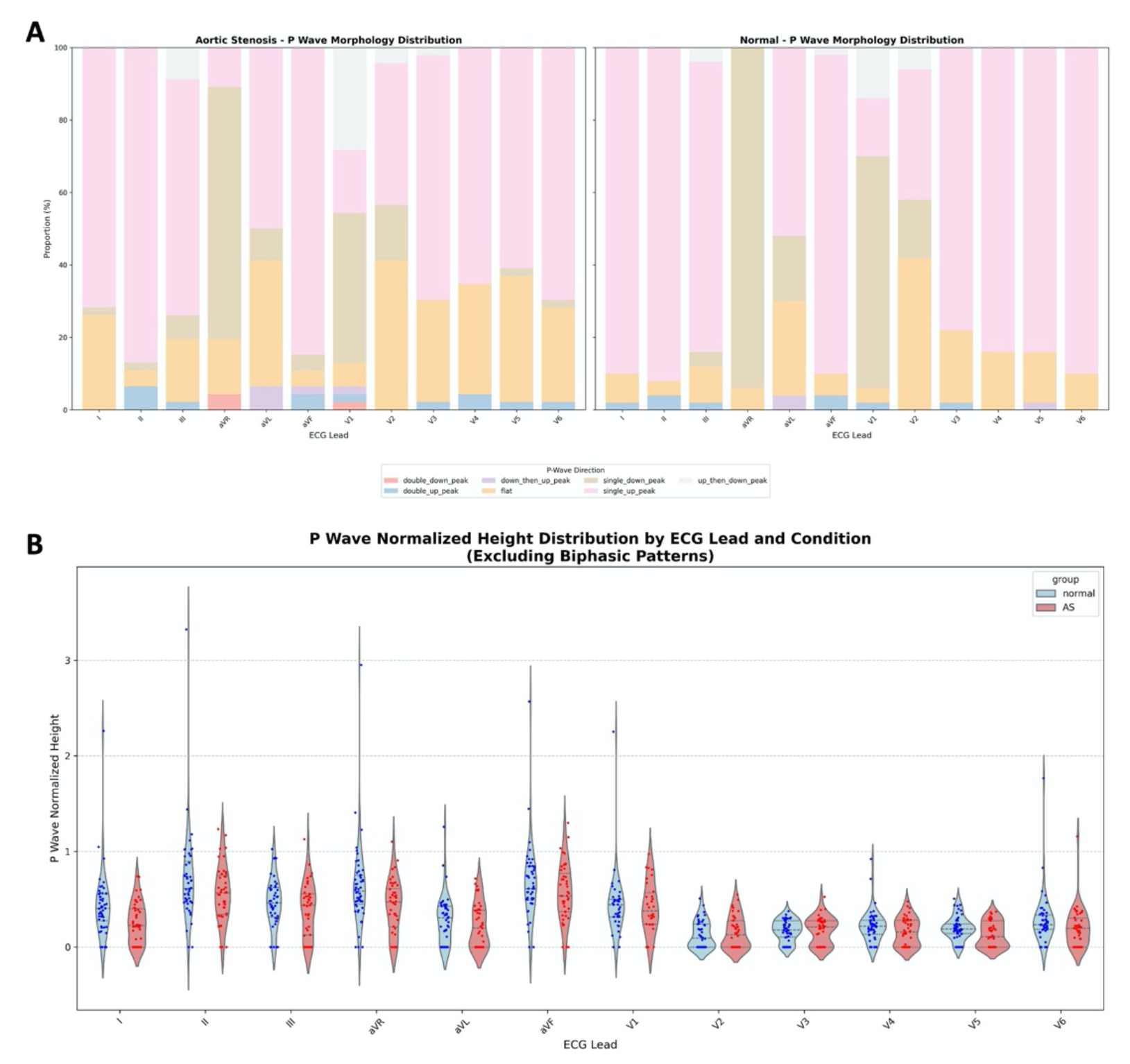
P-wave Morphology Distribution and P-wave Normalized Height Distribution for Normal ECG and ECG with aortic stenosis (AS) labeled in the associated ECHO report.

## DISCUSSION

In this study, we developed MERL-ECHO, the first CLIP-based multimodal learning framework that aligns 12-lead ECG signals with natural language ECHO reports using contrastive learning. MERL-ECHO allows accurate zero-shot classification of 10 SHD conditions, achieving an average AUROC of 0.69 in the internal test set and 0.72 in the external validation cohort, with the strongest performance in predicting left ventricular systolic impairment and dilation. Importantly, saliency mapping revealed clinically meaningful ECG features contributing to predictions, highlighting the model’s interpretability. Overall, MERL-ECHO shows strong potential as a scalable, generalizable framework for detecting structural heart diseases from ECGs without the need for labelled training data.

Previous work in ECG classification has largely relied on single-modality models using self-supervised or supervised learning. Neural networks such as RNNs and CNNs ^32–34^ demonstrated the ability to learn ECG feature representations and reduce the need for handcrafted rules. However, these models remain constrained by their design: they only utilize ECG signals, require extensive manual labelling of diagnoses from electronic health records (EHRs), and are limited to fixed label sets. As a result, they overlook the abundant contextual knowledge embedded in natural language clinical text and cannot naturally extend beyond predefined outputs.

In contrast, CLIP provides a more flexible and scalable paradigm. Unlike conventional neural networks that require fixed, manually curated label sets and labour-intensive relabelling for each new task, CLIP-based architectures directly align ECG signals with free-text reports already available in electronic health records. This removes the bottleneck of extensive expert annotation, since the natural language text itself provides supervision. As a result, new diagnostic concepts can be incorporated simply by adding text prompts, without retraining the model. To our knowledge, this is the first demonstration of leveraging ECHO natural language text report within a multimodal CLIP model to link ECG with echocardiographic findings.

The CLIP architecture utilized contrastive learning methods to learn the relationship between ECG and ECHO text reports with two input encoders: an ECG encoder and a text encoder. Our experiments revealed a stark contrast in how different ECG encoder architectures respond to increased training data. The CNN-based ResNet18 model with ECG-ECG text pretraining demonstrated consistent and steady performance gains as the dataset size grew, with its AUROC increasing by 9% when scaling from 20% to 100% of the training data. Notably, with ECG-ECG text pretraining, using ResNet18 as the ECG encoder results in a higher AUROC of 7% than using ViT-Tiny as the ECG encoder; the reason behind this may stem from the different processing methods. CNN-based ResNet18 processes ECG digital signals through convolutional layers, which are good at extracting important local features from time-series data like ECG digital signals. Transformer-based Vit-Tiny does not have such an architecture and may struggle to extract local information. The continuous upward trend suggests that its performance has not yet reached saturation and could be further enhanced with additional data. This highlights the potential of training a large-scale foundation model on substantially larger ECG-ECHO datasets, which could enable more robust zero-shot predictions across an even broader range of cardiac conditions.

Saliency mapping confirmed expected predictors of left ventricular impairment, including abnormal R-wave progression and ventricular ectopy, consistent with existing clinical understanding. Unexpectedly, the model consistently highlighted the P wave as most important for predicting aortic stenosis. Further analysis showed a higher prevalence of flattened P waves in leads I, V5, and V6 among affected patients. Previous ECG studies in aortic stenosis have mainly focused on QRS complex, ST segment and T wave^35–38^. Atrial findings have usually been limited to changes in P-wave duration and dispersion ^39,40^. Subtle P-wave flattening has not been systematically reported. These findings suggest that atrial remodeling in aortic stenosis may be detectable through P-wave. Independent validation and mechanistic studies are needed to clarify whether this feature provides additional diagnostic or prognostic value.

One possible future development of CLIP architecture for ECG classification is to expand the multi-modal training to ECHO videos, as the raw spatial information may also provide useful features for training ECG classifiers. However, it is well recognized that ECHO videos typically require multi-frame sampling or temporal modeling ^41,42^, which increases inference time and GPU memory pressure. It is therefore technical challenging to train such a model at the moment.

### Limitations

Our model has several limitations that warrant consideration. First, the text encoder used in this study was primarily trained on English text. ECHO reports written in other languages would require a language-specific encoder to ensure reliable performance. Second, as this is a retrospective study, a prospective clinical trial is necessary to assess the model’s accuracy and generalizability in real-world clinical settings. Third, the presence of class imbalance in our datasets may have contributed to overly optimistic AUROC values for certain minority classes during testing.

## CONCLUSION

MERL-ECHO demonstrates that aligning ECG signals with natural language echocardiography reports via contrastive learning enables robust, interpretable, and generalizable zero-shot detection of structural heart disease.

## ABBREVIATIONS

AUPRC: Area under the precision-recall curve
AUROC: Area under the receiver-operating characteristic curve
CNN: Convolutional neural network
DNN: Deep neural network
ECG: Electrocardiography
ECHO: Echocardiography
Grad-CAM: Gradient-weighted class activation mapping
RNN: Recurrent neural network
SHD: Structural heart disease

## Author Contributions

WCW, CL and CKW conceived the study design. WCW, PE, JWH, CYL, XYQ, HLL, YML, AC, CHY, CHF, WKC, CKC, LLC, LML, RR, JQ, HOACC and CKW acquired internal and external data. WCW, CL, XYQ, YML, CFT, RR, JQ, LY, HWL, RA, JWKH and CKW contributed to machine learning model development and statistical analysis. CYL, HHL, YML, HOACC, HFT, CWS and CKW provided clinical expertise for the study. WC, YML and CKW wrote first draft of the manuscript. CL, CYL, HOACC, HFT and CWS revised the manuscript for intellectually critical content. All authors read the final manuscript and authorized its submission.

## Data Availability Statement

Pre-training data (ECG-ECG text data) are all publicly accessible datasets from MIMIC-ECG ^23^. Fune-tuning data (ECG-ECHO text data) at Queen Mary Hospital and Tung Wah Hospital will not be publicly available owing to institutional policy. External test data from Columbia are available on PhysioNet https://physionet.org/content/echonext/1.0.0/. Codes and model weights are available at https://github.com/ConstantineWong/MERL-ECHO. Other anonymized data are available at reasonable request from the corresponding author for 3 years from date of publication.

## Sources of Funding

The study was partly supported by Rosie Young Medical Fellowship for Internal Medicine, Sun Chief Yeh Heart Foundation, Hong Kong.

## Acknowledgement

None.

## Disclosures

The authors report no conflict of interest.

## Notes

**Disclosure:** The authors report no conflict of interest.

### Competing Interest Statement

The authors have declared no competing interest.

### Author Declarations

Ethical approval was granted by HKU/HA HKW Institutional Review Board (HKU/HA HKW IRB; UW 25-042).

